# Whole population cohorts versus sampled comparators designs for evaluating health and educational outcomes of children with inborn rare conditions: a simulation study

**DOI:** 10.64898/2026.01.09.26343758

**Authors:** Joachim Tan, Milagros Ruiz Nishiki, Mario Cortina-Borja, Rachel L Knowles, Katie Harron, Catherine Peters, Pia Hardelid

## Abstract

**Background:** Linked administrative data covering whole populations are fundamental resources for longitudinal studies of children with rare conditions (cases) and unaffected peers (comparators). Data minimisation regulations sometimes limit the number of comparators per case (sampled comparators, ***SC***), with unknown impact on study findings.

**Methods:** Using Monte Carlo draws, we simulated 100 000 children with and without an exemplar condition, congenital hypothyroidism (CHT), with covariates (sex and comorbidity). Three outcomes (*Y*: Maths tests *z*-score, age 11 years; *L*: achieving expected Maths attainment (binary); *T*: months to neurodevelopmental disorder diagnosis) were modelled as linear combinations of CHT, sex and comorbidity. Varying parameters (comorbidity prevalence; comorbidity-CHT association; CHT effect on *Y/L/T*) factorially produced 36 data-generating mechanisms (DGMs). We used regression coefficients (CHT effect), standard errors (SEs) and *p*-values from 1000 simulations to evaluate power, precision and bias, comparing ***SC***n (n=5/10/15/25/50/100) versus full cohort (***FC***).

**Results:** Mean *p-*values and SEs for ***SC***25 generally deviated ≤5% versus ***FC*** with medium effects (*z*-score difference=0.3; odds/hazard ratios≥2), and ≤2% for large effects (*z*-score difference≥0.6; odds/hazard ratios≥5). For all outcomes, no ***SC*** nor ***FC*** had sufficient power (>80% of *p*-values≤0.05) with small or medium effects, whilst all ***SC*** had sufficient power with large effects. Compared with ***FC***, precision loss for ***SC***25 was 2.0-4.3%, 5.0-8.9%, 6.7-15.5% for *Y*, *L*, *T* respectively. ***SC*** was not associated with bias.

**Conclusion:** ***SC***25 provided comparable performance as ***FC*** for rare disease studies under several scenarios, but small effects posed challenges, notwithstanding sampling. This approach generates cost-effective recommendations for study design and data minimisation.

**Key messages:** What is the minimum ratio of children without disease (comparators) to children with disease (cases) needed to reliably quantify differences in health and educational outcomes, if whole population data were not accessible?

Sampling 25 comparators per case would generally provide comparable inferences as whole population data for typical scenarios likely to be encountered in longitudinal studies involving children with rare diseases.

Decreasing sample sizes helps studies to fulfil data minimisation principles, guides negotiations with data providers and facilitates approvals by research governance bodies, without compromising the quality of research findings.

## Background

Improved treatments and neonatal care have contributed to the increased survival of children with inborn conditions, such as congenital malformations and inherited disorders. Many inborn conditions are rare, affecting <5 per 10 000 individuals,[1,2] and lack effective treatments. These children are more likely to need health, social and educational support services than their peers.[3,4] Parents report having to manage uncertainty about their child’s prognosis,[5,6] alongside health professionals’ unfamiliarity with wider care needs.[7] The 2013 UK Strategy for Rare Diseases and successive policy documents stressed the lack of longitudinal, population-based data to assess the aetiology and health outcomes of rare conditions to support parents, clinicians and commissioners.[1,2,8] Assessing the long-term health and education outcomes for children with these conditions is a priority.

Disease registries, such as the National Congenital Anomaly and Rare Disease Registration Service,[9] contain detailed data necessary for case ascertainment and surveillance, but they seldom include long-term (>1 year) follow-up information due to the substantial resources involved. Moreover, many registries only collect data on children with a particular condition,[10] and health or developmental outcomes cannot easily be compared with their unaffected peers or other groups. Furthermore, information on potential confounding, mediating or modifying factors, such as comorbidities or socioeconomic status, may be absent.

Linkage between clinically validated registry data and routinely collected administrative health, education and social care records offers an efficient (and sometimes the only) method of following-up children with rare diseases (hereafter cases). It also enables comparisons with their unaffected peers from the same population (comparators) within the administrative data; we eschew the term ‘controls’ to avoid confusion with case-control studies. Successful examples of such studies are available from the UK, across Europe, and internationally.[11–17] Some studies have access to data for all comparators in the population (full cohort), whilst others will use randomly sampled comparators. The latter tend to be favoured by governance and ethics committees because of non-disclosure and data minimisation considerations, but may fail to produce samples that reflect the true distribution of important confounding comorbidities when these are prevalent amongst cases, yet are uncommon in the general population.[18–20]

We aimed to compare the statistical inferences generated under sampled comparators versus full cohort study designs for investigating rare disease outcomes. Our motivating example was evaluating the health and education outcomes for children with congenital hypothyroidism (CHT), an inborn endocrine disorder that affects 3-4 per 10 000 livebirths in Europe.[21] A data governance committee overseeing sharing of confidential patient data for research previously recommended using sampled comparators instead of a full cohort.[22] We sought to demonstrate the circumstances under which sampled comparators could produce findings of similar quality to full cohorts whilst achieving data minimisation.

## Methods

### Study design

We used Monte Carlo draws to generate schematic datasets containing exposure, outcome and confounder variables, guided by parameters extracted from official statistics and published studies. Regression models were fitted to estimate the association between CHT and outcomes, using full cohort and different sampled comparator designs, under multiple scenarios (reflecting different assumptions about disease prevalence, strength of association, and confounders’ influence). These scenarios correspond to data-generating mechanisms within the framework of Morris et al.[23]

#### Aims

To investigate how varying the number of sampled comparators per case (SC) would impact analytical estimates, with a view to determining the optimal SC to inform future study designs.

#### Data-generating mechanisms (DGMs)

We simulated datasets containing 100 000 children, approximately the annual number of births in the North Thames region – incorporating 24 North London local authorities and parts of Essex, Hertfordshire and Bedfordshire.[24] This is the catchment area of a large regional newborn blood spot screening programme for ten inborn conditions, including CHT; since 2006, babies who screened positive for CHT would be referred to Great Ormond Street Hospital for diagnosis and treatment.[25] Data from these sources were used to inform our parameters for sex-specific CHT prevalence.

##### Exposures

The dataset contained CHT status (yes/no), sex (male/female) and comorbidity status (yes/no), as well as selected outcomes of interest (see below). Sex was generated as a Bernoulli variable with an individual’s probability of being male (π_MALE_)=0.5124, based on the sex distribution using mid-year population estimates for age <1 year (averaged over 2002-2019) for the local authorities which make up the North Thames region, published online by the Office for National Statistics.[26] Comorbidity functioned as a placeholder confounder of the association between CHT and educational outcome. We used Down Syndrome as an exemplar comorbidity, because the prevalence of CHT has been estimated to be 28-35 times higher amongst children with Down syndrome than in the general population.[27] The probability of the *i*th individual being a CHT case, as a function of comorbidity (CMB hereafter) and sex, is given by equation (1) (see box).

**Data model equations and performance measures**

**Equations**

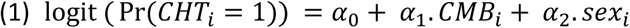

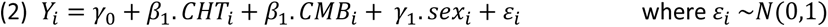

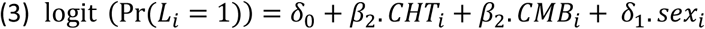

CHT: congenital hypothyroidism; CMB: comorbidity

Values of coefficients are given in Table 1 and Table 2 below.

**Performance measures**

i) Power: probability of detecting a difference (where it exists), measured by the proportion of *p*-values ≤0.05 from *z*-tests of *β̂*. We took ≥80% power at the 5% level of significance to be the adequacy threshold.
ii) Mean of model-based SE of the estimates, *β̂*
iii) Relative precision: derived from the variances of *β̂* under each SC compared against FC

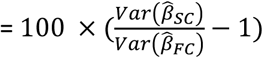

iv) Bias: mean of the distance between *β̂* and *β*

Formulas for each measure can be found in Morris et al.[23]

The order of derivation created an association between CHT and CMB, whilst remaining agnostic regarding the direction or mechanism of causation. Values for the model’s parameters *α*_0_*, α*_1_ and *α*_2_ were chosen using published CHT prevalence and hospital referral data, and studies of Down syndrome and CHT (Table 1).[24,28]

**Table 1:** Summary of parameters used for data generation.

| Description of data model attributes | Associated parameter | Constant or variable | Value(s) |
| --- | --- | --- | --- |
| **Baseline characteristics** |  |  |  |
| Probability of being male | $\pi_{\text{MALE}}$ | Constant | 0.5124 |
| Prevalence of comorbidity (CMB), proportion of population | $\pi_{\text{CMB}}$ | Variable | 0.0002, 0.001, 0.01, 0.05 |
| Base probability of being a CHT case | $\exp(\alpha_0)$ | Constant | 0.004 |
| Relative prevalence of CHT in those with and without CMB ($R_{\text{CMB}}$) | $\exp(\alpha_1)$ | Variable | 5×, 10×, 30× higher |
| Prevalence of CHT in males compared with females | $\exp(\alpha_2)$ | Constant | 2× lower |
| **Outcomes** |  |  |  |
| *Y*: Standardised score in Maths test at age 11 |  |  |  |
| Base mean score (intercept) | $\gamma_0$ | constant | -0.04 |
| Mean difference in *Y*, males vs. females | $\gamma_1$ | constant | 0.08 |
| Mean difference in *Y*, CHT vs. non-CHT | $\beta_1$ | variable | see Table 2 |
| *L*: Achieving expected attainment in Maths at age 11 |  |  |  |
| Base log odds of *L* | $\delta_0$ | constant | 1.277 |
| Log odds ratio, males vs. females | $\delta_1$ | constant | -0.041 |
| Log odds ratio, CHT vs. non-CHT | $\beta_2$ | variable | see Table 2 |
| *T*: Time to diagnosis of developmental disorders, years |  |  |  |
| Log hazard ratio, CHT vs. non-CHT | $\beta_3$ | variable | see Table 2 |
| Scale parameter | $\lambda$ | constant | 0.005 |
| Shape parameter | $\kappa$ | constant | 0.9 |

##### Outcomes

Three exemplar outcomes, representing health and educational endpoints, were generated using different data distributions: (*Y*) representing standardised marks in Maths tests sat by pupils at age 11 in England (normally-distributed); (*L*) achieving the expected level of attainment in Maths at age 11, based on teacher-assessments (binary)[29]; (*T*) age at diagnosis of a neurodevelopmental disorder (time to event). Equations for *Y* (2) and *L* (3) are shown in the box.

As contemporary studies of the educational attainment of children with CHT are lacking, we posited that attainment differentials (*β*_1_ and *β*_2_) between children with and without CHT might be broadly similar, on average, to those reported elsewhere for children with and without major congenital anomalies.[30–32] We used coefficients from different anomaly subgroups to exemplify three levels of attainment deficit based on their likely developmental impact: small (hypospadias, syndactyly), medium (congenital heart defects, cleft palate) and large (congenital hydrocephalus). This allowed us to model degrees of cognitive impairment, from mild to substantial, reflecting CHT severity and treatment status. As we always adjusted for CMB in our models, the effect of CMB on outcome was immaterial; we set it to that of CHT to minimise the number of distinct parameters.

*T* was modelled using a Weibull distribution with scale (*λ*) and shape parameters (*κ*) and log hazard ratio *β*_3_ for CHT versus non-CHT. Values were chosen to approximate the cumulative incidence of specific developmental disorders of motor function, scholastic skills, speech and language at age 15 years in CHT cases compared with controls, reported elsewhere.[33]

##### Varying parameters

The probability of CMB (π_CMB_) was varied: 1 in 5000, 1 in 1000, 1 in 100, 1 in 20. We also varied *α*_1_ such that the probability of having CHT was 5, 10 and 30 times higher for those with CMB versus those without (relative prevalence, R_CMB_), which indirectly changed CHT prevalence. We chose three values for each parameter *β*_1,_ *β*_2_ and *β*_3_ to represent a weak, moderate and strong association between CHT and outcome; for convenience, we shall refer to them interchangeably as effect sizes (small, medium and large respectively), without implying a causal mechanism. DGM parameters are summarised in Table 1 and Table 2.

**Table 2:** Outcome measures and parametrisation of effect sizes, comparing CHT vs. non-CHT.

| Outcome | Measure | parameter | Effect size category and parameter values |  |  |
| --- | --- | --- | --- | --- | --- |
|  |  |  | small | medium | large |
| *Y* | Mean difference | $\beta_1$ | -0.03 | -0.3 | -0.6 |
| *L* | Odds ratio |  | 0.92 | 0.48 | 0.17 |
| | log odds ratio | $\beta_2$ | -0.08 | -0.74 | -1.78 |
| *T* | Hazard ratio |  | 1.5 | 3.0 | 5.0 |
| | log hazard ratio | $\beta_3$ | 0.41 | 1.10 | 1.61 |

#### Estimands

The estimands *β*_1_, *β*_2_ and *β*_3_ (collectively denoted by vector ***β***) represented the effect of CHT (versus no-CHT) on *Y*, *L* and *T* respectively.

#### Methods for simulation and analysis

For each of 36 DGMs [π_CMB_(4) × R_CMB_(3) × effect(3)], we generated 1000 simulated datasets. Datasets were analysed using linear, logistic, and Cox proportional hazards regression models (adjusting for sex and CMB) to obtain estimates of ***β***, standard errors (SEs) and *p*-values. Data were first analysed with full cohort (FC), then by subsets containing all cases and a random sample of *n* comparators per case (abbreviated to SC*n* hereafter, where *n=*5/10/15/25/50/100). Estimates from each SC and FC were aggregated over 1000 simulations for each DGM.

Performance Measures (see box)

### Statistical analysis

We used summary statistics and visualisations to check that the generated data conformed to the parameters specified by the respective DGMs. Regression estimates ***β̂*** were examined for model convergence (within 100 iterations) and missing or uninterpretable estimates (e.g. infinitely large absolute hazard ratios, usually from perfect separation) were excluded. Comparing different SC versus FC, we considered a departure of ≤2% to be non-inferior to FC and >2% and ≤5% to be satisfactory for every performance measure. We examined Monte Carlo errors which quantified the uncertainty of performance measures from a finite number of simulations. All simulations and analyses were performed in Stata version 18.5 (StataCorp LLC, Texas, USA). Survival times *T* were generated with the *survsim* package,[34] and we used *siman* (incorporating lolly and nested-loop plots) for performance evaluation.[35,36]

## Results

We obtained 252 000 sets of results from all DGM and SC combinations over 1000 simulations each. Figure 1 shows CHT prevalence varying by π_CMB_ and R_CMB_; across all DGMs the mean was 3.6 (range: 0.9, 10.2) per 10,000 population. Generally, there were few individuals with both CHT and CMB (median [IQR]: 0 [0–0], 0 [0–1], 3 [1–7], 15 [9–40] for π_CMB_=0.0002, 0.001, 0.01, 0.05 respectively.)

**Figure 1:**
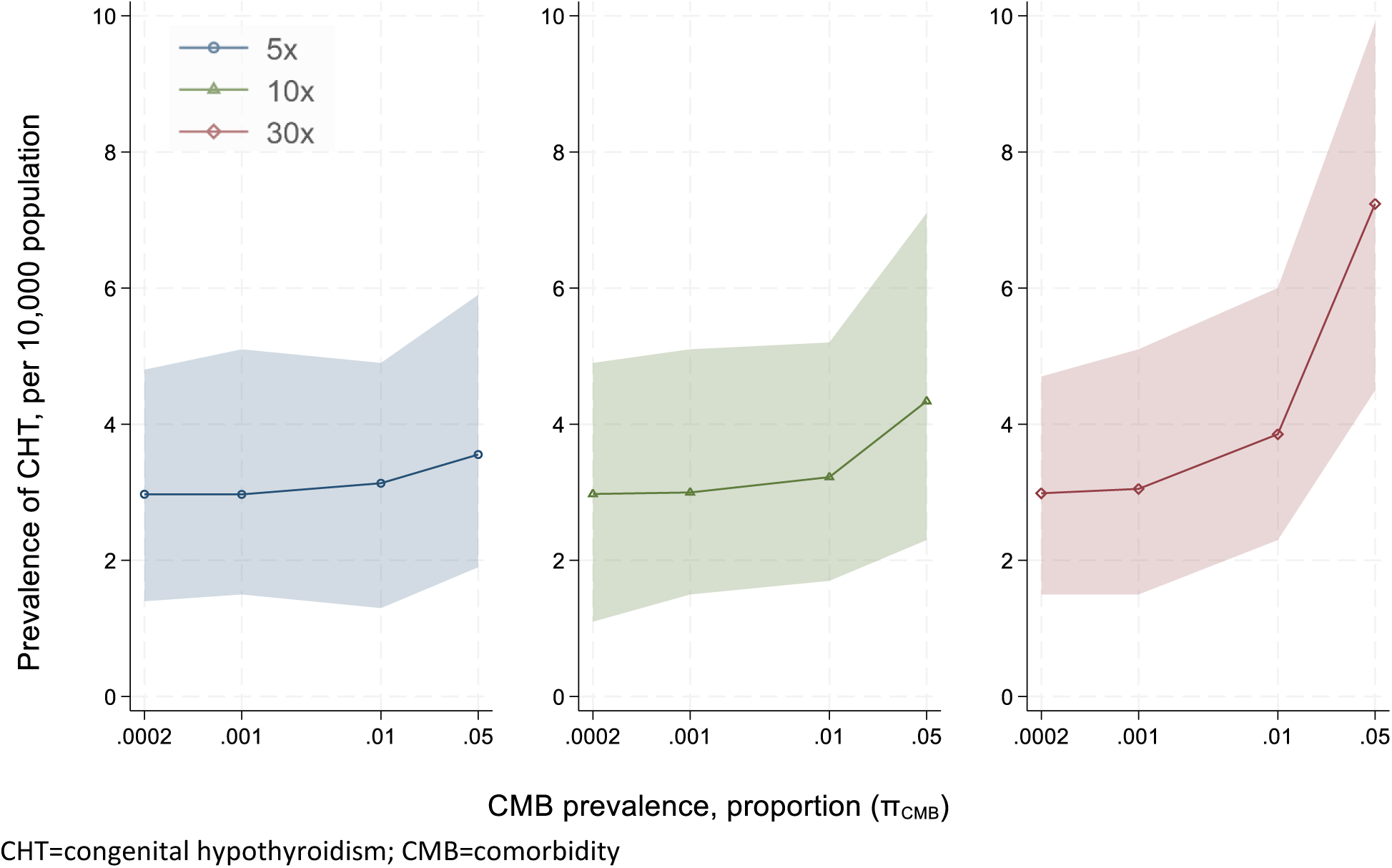
Prevalence of children with congenital hypothyroidism (CHT) per 10 000 population, versus proportion of comorbidity (π_CMB_), by levels of R_CMB_ (Left=5×; Centre=10×; Right=30×). Shaded areas indicate range from 1000 simulations. CHT=congenital hypothyroidism; CMB=comorbidity

Figure 2 shows effect measures for *Y* and *L*, comparing CHT and non-CHT children, from 1000 simulations under one DGM (π_CMB_=0.01, R_CMB_=10x); the distances between observed means and true parameter values were within expectation.

**Figure 2:**
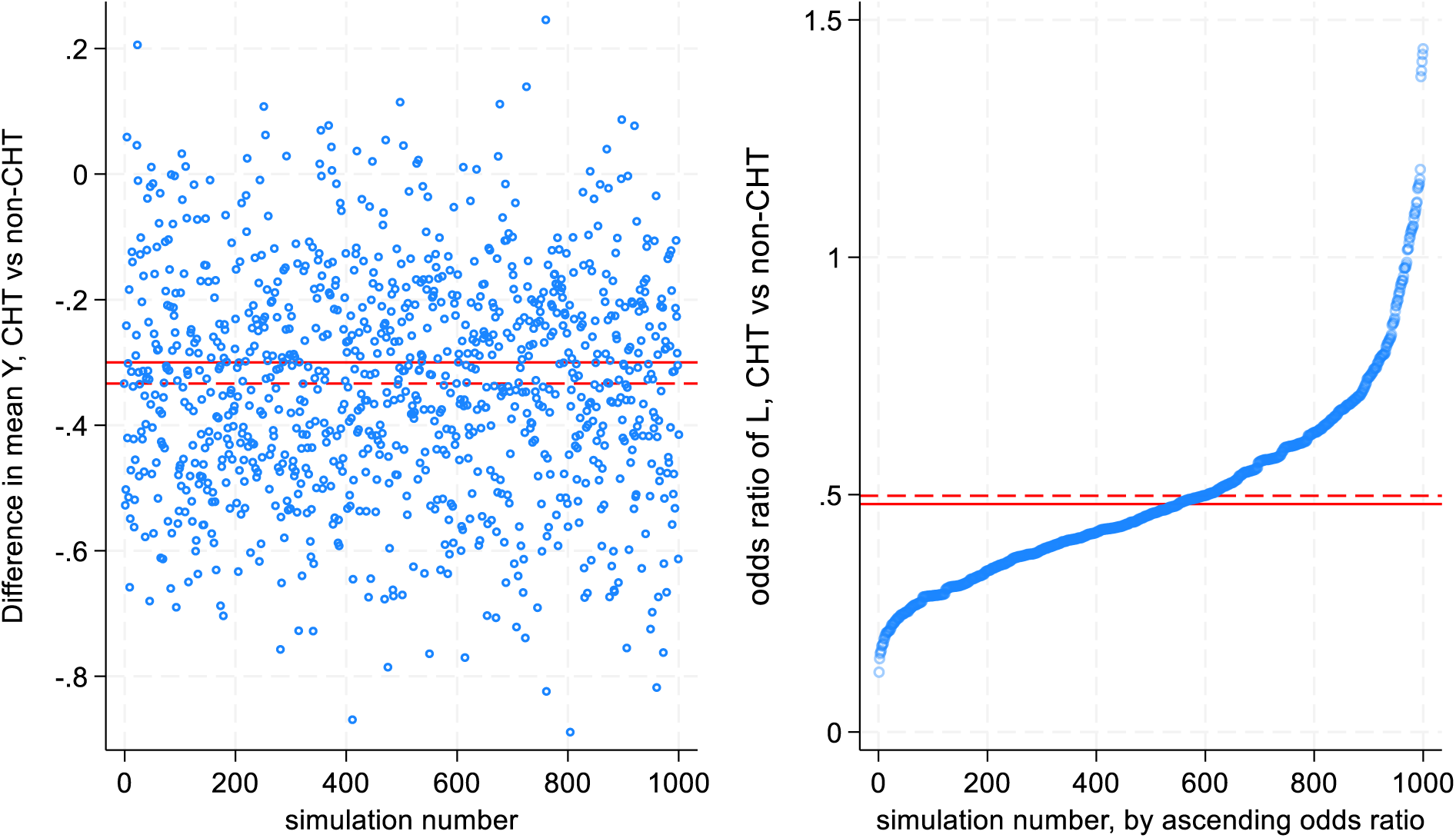
Difference in mean *Y* (left) and odds ratio for *L* (right) from 1000 simulated datasets, comparing CHT vs. non-CHT, when π_CMB_=0.01, R_CMB_=10x. Dashed lines indicate mean value from 1000 simulations; solid lines represent true parameter.

### Performance Measures

#### Power

For *Y*, Figure 3 illustrates how power varied with SC under each DGM. When the effect size (*β*_1_) was small or medium, none of the SC (including FC) had adequate power, but for medium effects, there was visible separation between different SC, increasing with the number of comparators. With large effects, SC25 seemed similar to FC for almost all DGMs. Table S1 shows exact values and Monte Carlo errors for a subset of DGMs when R_CMB_=10×; SC25 was satisfactory for medium effects when π_CMB_ ≤0.001, and was mostly non-inferior to FC for large effects.

**Figure 3:**
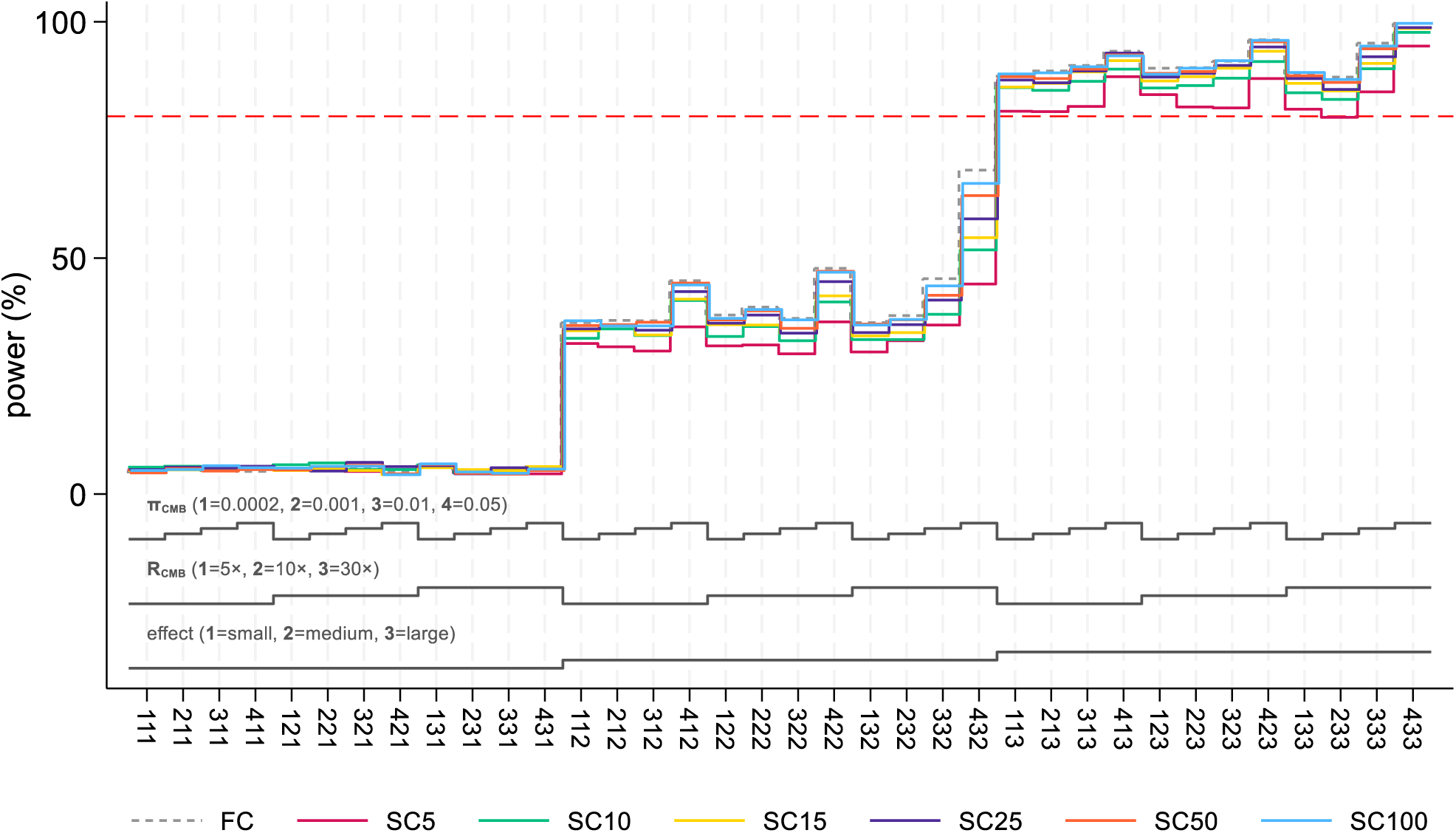
A nested-loop plot of power for *Y* comparing CHT vs. non-CHT, under different SC and data-generating mechanisms. Three-digit numbers on the x-axis represent unique ordered combinations of π_CMB_, R_CMB_ and effect size *β*_1_ (each digit is a code for the corresponding value of the parameter as shown below). Red dashed line denotes power=80%.

For *L*, there was almost no differential between SC for small and large effects. For medium effect, the gap between SC25 and FC widened with higher π_CMB_ and R_CMB_, making SC50 or SC100 a better choice for approximating FC (Figure S2a). With *T*, power with SC25 seemed reasonably close to FC for medium and large effects; when effect was large, SC5 also met the threshold for adequate power for most scenarios (Figure S3a and Table S4).

#### Model standard errors (SE)

Model SE decreased with increasing π_CMB_, R_CMB_, and SC, as all contributed to larger sample sizes available for analysis (Figure 4). R_CMB_ exerted a greater influence on model SE when π_CMB_ ≥0.001, mirroring the pattern of increasing CHT cases (Figure 1). When R_CMB_=30× there was increasing separation with higher π_CMB_ between different SC (fanning out) for *Y* and *L*.

**Figure 4:**
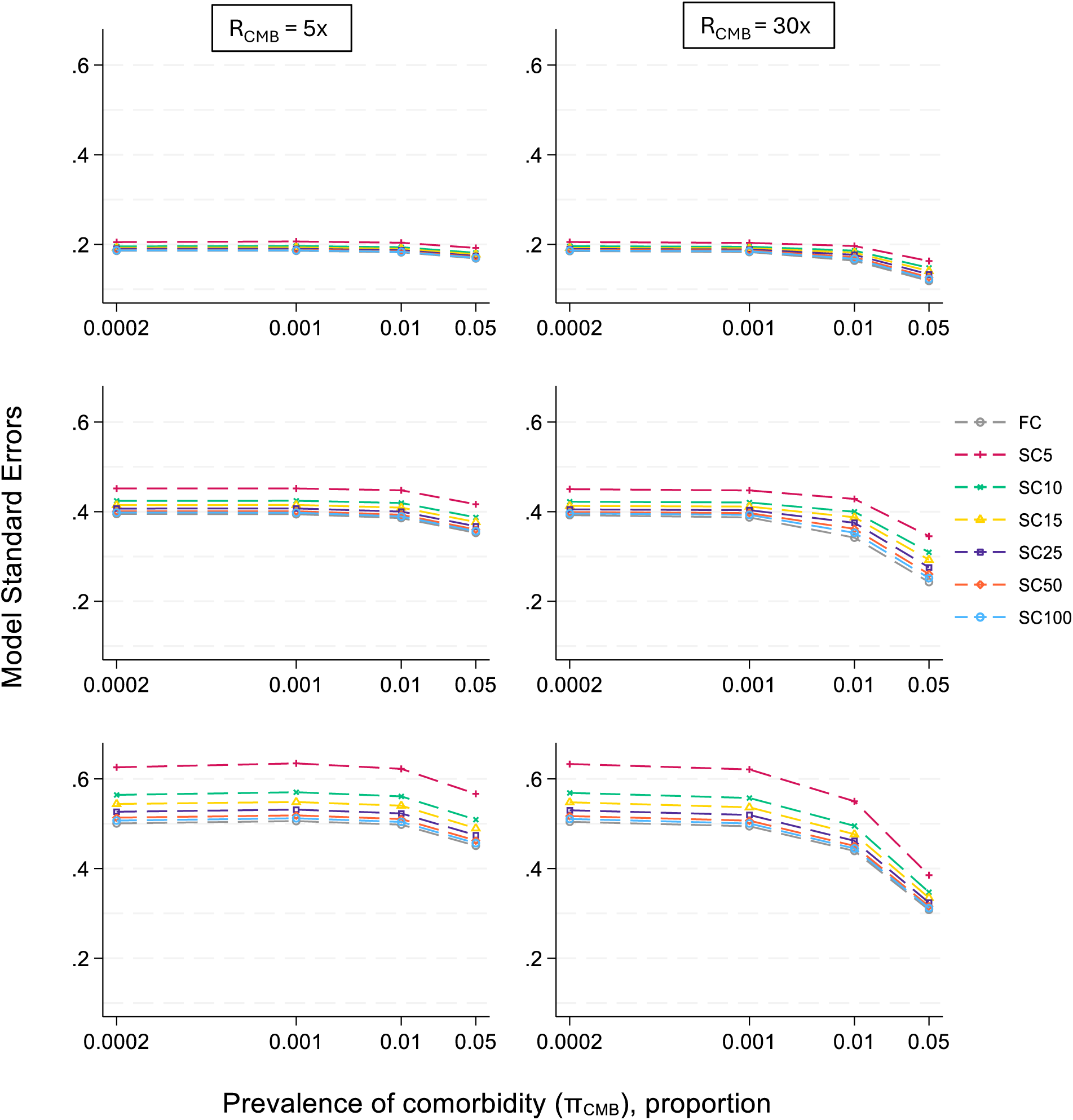
Model standard errors of estimates *β̂*_1_, *β̂*_2_and *β̂*_3_ for *Y* (top), *L* (middle) and *T* (bottom) versus π_CMB_, when R_CMB_ = 5× and 30×, and effect size is medium.

For most DGMs, model SE under SC25 differed from FC by ≤5% (Figure 5), except when R_CMB_=30× (given considerably smaller initial values of model SEs under FC). With low values of π_CMB_ and R_CMB_, SC25 generally provided a good approximation to FC, whilst SC5 was distinctly further from FC than other SC throughout. Model SEs did not vary significantly across effect sizes (not shown).

**Figure 5:**
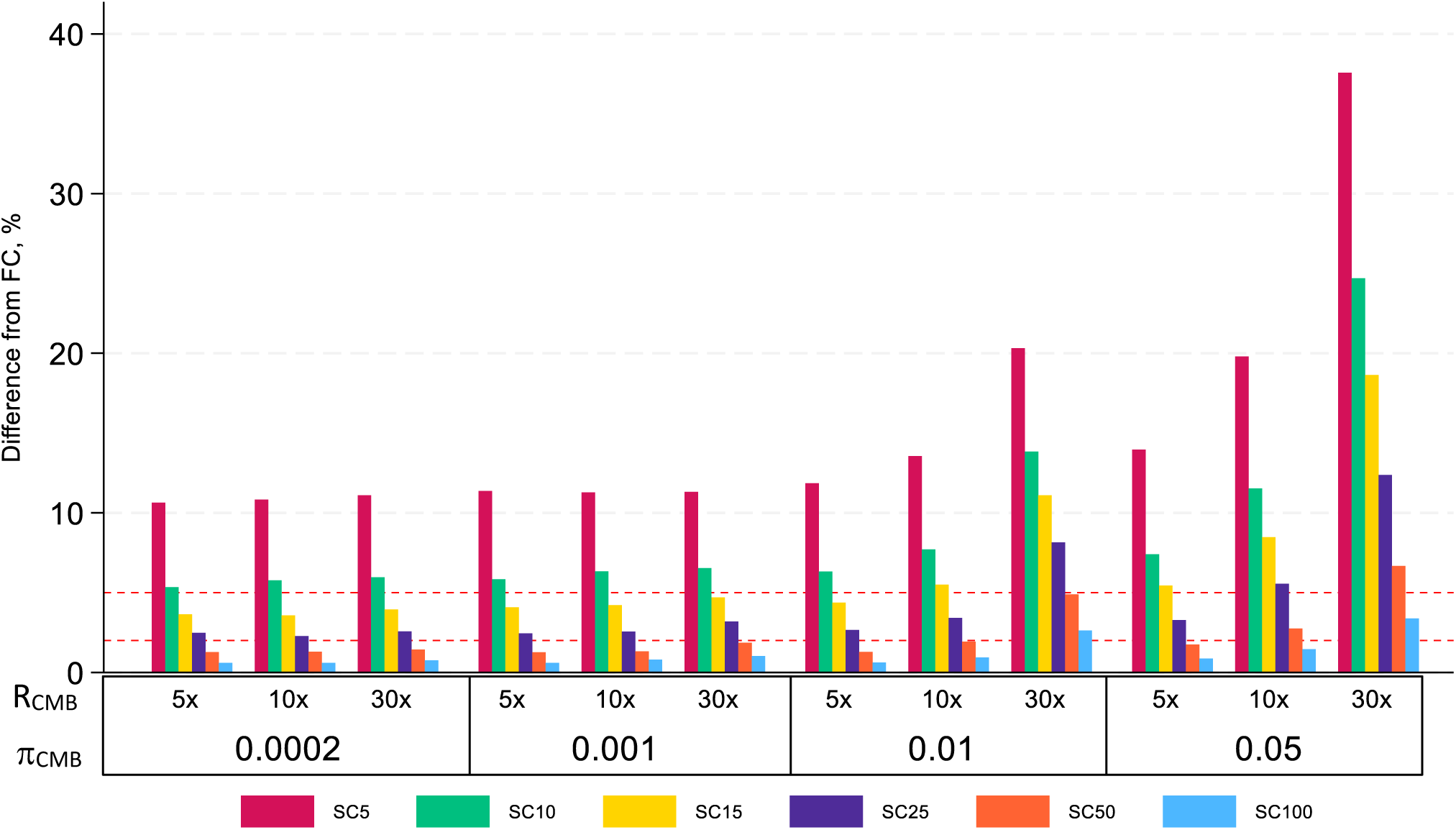
Percentage difference in model standard errors for *β̂*_1_ between different SC versus FC, by π_CMB_ and R_CMB_ when *β*_1_ =-0.3. Dashed lines represent 2% and 5% thresholds.

#### Relative precision

Relative to FC, all SC incurred a loss of precision (greater variation in ***β̂*** across simulations) that was inversely related to SC size (Figure 6). This occurred across all outcomes and effect sizes, with few exceptions (e.g. large effect, *Y* and *L*). Additionally, for *T*, the magnitude of precision loss increased with larger effect sizes.

**Figure 6:**
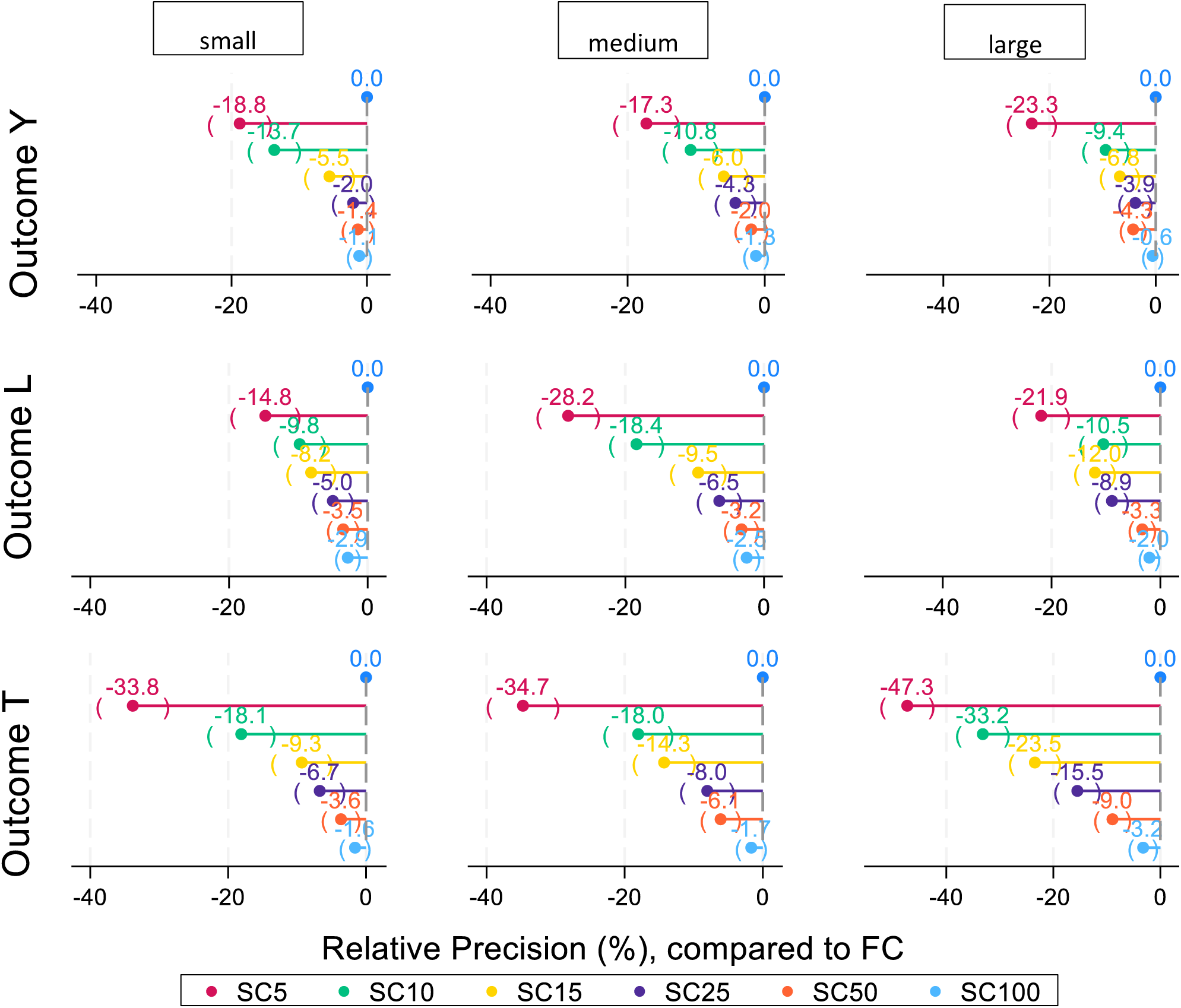
Relative precision (%) for different SC designs compared to FC for *Y*, *L* and *T*, by effect size when π_CMB_=0.001 and R_CMB_=10×. Negative value indicates precision loss; brackets show Monte Carlo errors.

#### Bias

There was no significant bias in estimating the true mean difference in *Y*, comparing CHT vs. non-CHT (Figure S1b). For *L*, there appeared to be systematic overestimation and underestimation of the true log odds ratio for small and large effects respectively, regardless of SC and DGM (Figure S2b).

Cox proportional hazards models tended to underestimate the true hazard ratio throughout, but there was no clear association between SC and bias; unexpectedly SC5 seemed to produce the least biased estimate across DGMs (Figure S3b).

### Monte Carlo errors

Monte Carlo errors showed similar patterns for all outcomes, and were lowest for model SE (<1% across almost all the DGMs), and did not vary appreciably by SC. Monte Carlo errors for relative precision, power and mean remained <5% generally, but exceeded 10% for power and mean in *Y* and *L*, when effect size was small (Tables S2, S3 and S4).

### Estimation problems

Of 252 000 results, 51 (0.02%) logistic regression models for *L* failed to produce an interpretable estimate (log odds ratio=0). All of these occurred with small effect size, and were not strongly influenced by π_CMB_ or SC. In contrast, estimation problems for *T* occurred 6093 (2.42%) times and were more frequent with smaller values of π_CMB_, R_CMB_ and effect size, and but not SC (Table S5).

## Discussion

Based on key performance measures, SC25 provided reasonable approximation to FC for a range of scenarios likely to be encountered in rare disease studies. However, small effect sizes (weak associations between CHT and outcome) posed challenges for reliable inference, even with relatively common exposures and full cohort data. Reducing SC was not associated with systematic bias.

### Strengths and limitations

We examined how sampling a modest number of comparators, in aid of data minimisation, would affect findings from longitudinal studies of rare disease cohorts using linked administrative data. We based starting parameter values on published reports and studies to generate data that would be plausible, as well as varied magnitudes factorially to reflect different assumptions about disease prevalences and effect sizes, potentially increasing the applicability of our findings to other conditions and outcomes. Our approach could be extended with the inclusion of additional confounders, such as sociodemographic indicators (e.g. birth year, ethnicity and deprivation) or multi-level data (e.g. clustering by regions) to help refine study design.

The finite number of DGMs and SC that we could simulate did limit granularity. For example, SC20 may have performed similarly to SC25 and would have better achieved data minimisation. Likewise, we were limited to examining only three categories of effect sizes. Increasing the number of DGMs would have greatly increased computational time, and we pragmatically selected a few values for each parameter to illustrate general features and uncover underlying patterns.

One difficulty was the lack of longitudinal studies on children with CHT (e.g. educational attainment in England) from which accurate effect sizes could be extracted. We have used a range of values reported for children with other conditions[30,32] to allow for variations in disease severity and inaccurate assumptions, albeit newborn screening and treatment greatly mitigate the risks of extreme cognitive sequelae. Nonetheless, our work underscores the need for wider research on the long-term outcomes of children with CHT.

### Interpretation

We found that overall, SC25 was broadly similar to FC in terms of preserving power and precision of estimates, across different types of study outcomes. This was consistent across a range of exposure and comorbidity prevalences, although gaps began to widen with increasing numbers of CHT cases, primarily due to more rapid gains in power and precision under FC. With very large effect sizes, even SC10 could be considered viable, depending on the chosen definition of satisfactory or non-inferior performance. However, accurate effect sizes, particularly for rare diseases, are unlikely to be known in advance.

Our study demonstrated that simulating data under simple parametric assumptions representing various scenarios is a feasible way of ascertaining lower limits for SC when there are data access constraints. These are useful for hypothesis generation or informing rules of thumb, and whilst we could further refine recommendations using more complex data, computational time increases exponentially with the number of DGMs, SC and analytical methods. Having 1000 repetitions might account for some of the relatively large Monte Carlo errors and anomalous patterns (e.g. worsening relative precision with more comparators) observed. A simpler data model which captures main features and could be run over vastly more repetitions may therefore be preferable, to ensure consistency and convergence.

Compared with continuous and binary outcomes, estimation of hazard ratios was more likely to fail with small effect sizes or low exposure/confounder prevalences. However, our value for a “small” hazard ratio (1.5) is not atypical for health outcomes. In such scenarios, the problems were attributable to the inherent sparsity of the data, which our choice of SC did not ameliorate or worsen. Increasing the sample size (e.g. including more years or regions) may be the sole solution.

A previous CHT study protocol proposed SC15 for investigating educational attainment, where it was hypothesised that differences between affected children and their peers would be smaller than 0.3 points in standardised scores.[22] That calculation was based on having access to multiple annual birth cohorts, containing a total of 1800 children with confirmed diagnosis of CHT. Our enquiry does not supplant sample size calculations, but examines what would be the lowest number of comparators which could provide similar inferences as FC for various outcomes under typical scenarios. Our findings suggest that SC25 would be broadly comparable to having FC data (notwithstanding that FC data may itself have insufficient power to answer a particular research question regarding outcomes for children with a rare disease).

This simulation study could be further developed to evaluate the impact of imperfect conditions characteristic of real-world studies, such as missing or misclassified data, and omission of confounders during analysis. In such cases, bias and coverage would be primary concerns, but loss of precision and power should also be quantified. Other extensions include using matched case-comparator designs with a view to potentially further reducing the number of comparators needed, and comparison of alternative methods for analysing hierarchical or panel data, commonly encountered in longitudinal studies of educational outcomes.

## Conclusion

Overall, having at least SC25 provided comparable performance to FC for rare diseases such as CHT under several scenarios, but small effect sizes posed distinct challenges regardless of sampling.

Where access to whole population data is not feasible, this approach provides a principled and cost-effective way to inform study design, achieve data minimisation and guide data negotiations.

Judicious selection of model parameters would enable extending this method for other diseases and outcomes.

## Ethics approval

Ethics approval was not needed for this study since it used no real data.

## Acknowledgements

We would like to thank Professor Emerita Bianca De Stavola for her advice and support for this project.

## Author contributions

PH and JT designed the study. MRN conducted the literature review. JT, PH and MCB developed the analytical strategy. JT wrote the Stata code, performed the analysis and drafted the paper. All authors contributed points to be included and critically revised the paper. JT is the guarantor.

## Supplementary data

Supplementary data are available at IJE online.

## Data availability

The data underlying this article have been generated by code which will be shared on reasonable request to the corresponding author. The *siman* suite of programs for analysing results of simulation studies is available from: https://github.com/UCL/siman.

## Use of Artificial intelligence (AI) tools

No AI tools were used.

