## Supplemental Material for "Whole population cohorts versus sampled comparators designs for evaluating health and educational outcomes of children with inborn rare conditions: a simulation study"

### Supplementary Tables

Table S1: Power to detect difference in Y between CHT and non-CHT of different SC, by true $\beta_1$ and $\pi_{CMB}$, holding $R_{CMB}$=10×. Bold and underscored numbers indicate ≤2.0% and >2.0-5.0% differences compared to FC, respectively. Shaded rows show Monte Carlo errors.

| $\beta_1$ (effect) | $\pi_{CMB}$ | *FC* | *SC5* | *SC10* | *SC15* | *SC25* | *SC50* | *SC100* |
|---|---|---|---|---|---|---|---|---|
| | 0.0002 | 5.5 | **5.4** | 6.2 | 5.0 | 5.2 | 5.1 | **5.5** |
| | | 0.721 | 0.715 | 0.763 | 0.689 | 0.702 | 0.696 | 0.721 |
| | 0.001 | 5.8 | 5.0 | 6.6 | 5.4 | 4.9 | **5.8** | **5.9** |
| **-0.03** | | 0.739 | 0.689 | 0.785 | 0.715 | 0.683 | 0.739 | 0.745 |
| **(small)** | 0.01 | 5.6 | 4.8 | 5.2 | 5.0 | 6.7 | 6.1 | 5.9 |
| | | 0.727 | 0.676 | 0.702 | 0.689 | 0.791 | 0.757 | 0.745 |
| | 0.05 | 4.8 | 5.2 | 5.2 | 4.1 | 5.8 | 4.3 | 4.1 |
| | | 0.676 | 0.702 | 0.702 | 0.627 | 0.739 | 0.641 | 0.627 |
| | 0.0002 | 37.9 | 31.4 | 33.4 | 35.9 | 36.2 | 36.9 | **37.2** |
| | | 1.534 | 1.468 | 1.491 | 1.517 | 1.520 | 1.526 | 1.528 |
| | 0.001 | 39.6 | 31.6 | 35.5 | 35.8 | 37.9 | **38.8** | **39.1** |
| **-0.3** | | 1.547 | 1.470 | 1.513 | 1.516 | 1.534 | 1.541 | 1.543 |
| **(medium)** | 0.01 | 37.2 | 29.7 | 32.5 | 34.1 | 34.1 | 35.1 | **36.9** |
| | | 1.528 | 1.445 | 1.481 | 1.499 | 1.499 | 1.509 | 1.526 |
| | 0.05 | 47.8 | 36.5 | 40.7 | 42.0 | 45.0 | **47.2** | **47.0** |
| | | 1.580 | 1.522 | 1.554 | 1.561 | 1.573 | 1.579 | 1.578 |
| | 0.0002 | 90.2 | 84.6 | 86.0 | 87.5 | 88.3 | **89.1** | **88.9** |
| | | 0.940 | 1.141 | 1.097 | 1.046 | 1.016 | 0.985 | 0.993 |
| | 0.001 | 90.3 | 82.0 | 86.5 | 88.4 | **89.1** | **89.5** | **90.2** |
| **-0.6** | | 0.936 | 1.215 | 1.081 | 1.013 | 0.985 | 0.969 | 0.940 |
| **(large)** | 0.01 | 91.7 | 81.8 | 88.1 | **90.2** | **90.8** | **91.8** | **91.8** |
| | | 0.872 | 1.220 | 1.024 | 0.940 | 0.914 | 0.868 | 0.868 |
| | 0.05 | 96.2 | 88.0 | 91.6 | 93.8 | **94.7** | **95.8** | **96.1** |
| | | 0.605 | 1.028 | 0.877 | 0.763 | 0.708 | 0.634 | 0.612 |

Table S2: Median values (range) of performance measures for outcome Y across all data generating mechanisms, for different sampled comparator (SC) versus full cohort (FC) designs, by effect size. Shaded rows show Monte Carlo standard errors of the median (expressed as %).

| Performance | Effect Size | SC5 | SC10 | SC15 | SC25 | SC50 | SC100 | FC |
|---|---|---|---|---|---|---|---|---|
| Power (%) | small | 5.1 (4.3, 5.9) | 5.2 (4.5, 6.6) | 5.1 (4.1, 5.8) | 5.6 (4.3, 6.7) | 5.1 (4.2, 6.3) | 5.4 (4.1, 6.4) | 5.3 (4.5, 6.2) |
| | | 13.6 | 13.5 | 13.6 | 13.0 | 13.7 | 13.2 | 13.3 |
| | medium | 31.8 (29.7, 44.5) | 34.3 (32.5, 51.7) | 35.7 (33.5, 54.3) | 36.1 (34.1, 58.3) | 36.9 (35.1, 63.2) | 37.1 (35.6, 65.8) | 37.8 (36.3, 68.6) |
| | | 4.6 | 4.4 | 4.2 | 4.2 | 4.1 | 4.1 | 4.0 |
| | large | 82.1 (79.8, 94.9) | 86.9 (83.6, 97.8) | 88.9 (85.4, 98.6) | 89.3 (85.7, 98.8) | 89.8 (87.2, 99.7) | 90.3 (87.8, 99.7) | 90.6 (88.3, 99.7) |
| | | 1.5 | 1.2 | 1.1 | 1.1 | 1.1 | 1.0 | 1.0 |
| Model SE | small | 0.21 (0.16, 0.21) | 0.20 (0.15, 0.20) | 0.19 (0.14, 0.19) | 0.19 (0.13, 0.19) | 0.19 (0.13, 0.19) | 0.19 (0.12, 0.19) | 0.18 (0.12, 0.19) |
| | | 0.4 | 0.3 | 0.3 | 0.3 | 0.3 | 0.3 | 0.3 |
| | medium | 0.20 (0.16, 0.21) | 0.20 (0.15, 0.20) | 0.19 (0.14, 0.19) | 0.19 (0.13, 0.19) | 0.19 (0.13, 0.19) | 0.19 (0.12, 0.19) | 0.18 (0.12, 0.19) |
| | | 0.3 | 0.3 | 0.3 | 0.3 | 0.3 | 0.3 | 0.3 |
| | large | 0.21 (0.16, 0.21) | 0.20 (0.15, 0.20) | 0.19 (0.14, 0.19) | 0.19 (0.13, 0.19) | 0.19 (0.13, 0.19) | 0.19 (0.12, 0.19) | 0.18 (0.12, 0.19) |
| | | 0.3 | 0.3 | 0.3 | 0.3 | 0.3 | 0.3 | 0.3 |
| Relative precision[1] (%) | small | -19.8 (-45.6, -17.3) | -12.7 (-36.3, -9.1) | -8.8 (-31.0, -5.3) | -6.5 (-20.9, -2.0) | -2.9 (-14.5, -1.4) | -1.9 (-7.3, -0.4) | -- |
| | | 2.2 | 2.2 | 2.2 | 2.2 | 2.2 | 2.2 | -- |
| | medium | -21.9 (-41.7, -16.4) | -13.2 (-31.0, -8.4) | -8.9 (-20.1, -4.9) | -5.2 (-18.7, -2.7) | -2.9 (-9.0, 0.2) | -1.1 (-5.7, 0.1) | -- |
| | | 2.2 | 2.2 | 2.2 | 2.2 | 2.2 | 2.2 | -- |
| | large | -23.7 (-48.2, -13.1) | -12.3 (-34.8, -9.4) | -9.3 (-30.7, -6.1) | -5.8 (-23.5, -1.7) | -3.2 (-10.0, -0.7) | -1.7 (-6.0, -0.6) | -- |
| | | 2.2 | 2.2 | 2.2 | 2.2 | 2.2 | 2.2 | -- |
| Mean point estimate | small | -0.03 (-0.04, -0.02) | -0.03 (-0.03, -0.02) | -0.03 (-0.03, -0.02) | -0.03 (-0.04, -0.02) | -0.03 (-0.04, -0.02) | -0.03 (-0.04, -0.02) | -0.03 (-0.04, -0.02) |
| | | -21.4 | -20.7 | -19.2 | -20.6 | -20.3 | -20.1 | -19.9 |
| | medium | -0.30 (-0.31, -0.29) | -0.30 (-0.31, -0.29) | -0.30 (-0.31, -0.29) | -0.30 (-0.31, -0.29) | -0.30 (-0.31, -0.29) | -0.30 (-0.31, -0.29) | -0.30 (-0.31, -0.29) |
| | | -2.1 | -2.0 | -2.0 | -1.9 | -1.9 | -1.9 | -1.9 |
| | large | -0.60 (-0.61, -0.59) | -0.60 (-0.61, -0.59) | -0.60 (-0.61, -0.59) | -0.60 (-0.61, -0.59) | -0.60 (-0.61, -0.59) | -0.60 (-0.61, -0.59) | -0.60 (-0.61, -0.59) |
| | | -1.1 | -1.0 | -1.0 | -1.0 | -1.0 | -1.0 | -1.0 |

1 Negative values represent precision loss. Monte Carlo errors shown are those of variance of $\beta_1$ (source from which Relative precision is calculated).

Table S3: Median values (range) of performance measures for outcome L across all data generating mechanisms, for different sampled comparator (SC) versus full cohort (FC) designs, by effect size. Shaded rows show Monte Carlo standard errors of the median (expressed as %).

| Performance Measure | Effect Size | SC5 | SC10 | SC15 | SC25 | SC50 | SC100 | FC |
|---|---|---|---|---|---|---|---|---|
| Power (%) | small | 5.1 (4.0, 6.8) | 5.0 (4.2, 6.1) | 5.3 (3.8, 6.1) | 5.2 (3.8, 6.4) | 5.2 (4.1, 6.9) | 5.4 (4.3, 7.4) | 5.2 (4.0, 6.4) |
| | | 13.6 | 13.8 | 13.4 | 13.5 | 13.6 | 13.2 | 13.5 |
| | medium | 41.6 (38.0, 59.2) | 44.5 (41.7, 66.2) | 46.6 (43.9, 71.3) | 48.5 (44.9, 77.2) | 49.3 (46.1, 80.6) | 49.6 (46.9, 82.5) | 50.5 (47.6, 86.1) |
| | | 3.7 | 3.5 | 3.4 | 3.2 | 3.2 | 3.2 | 3.1 |
| | large | 98.8 (97.8, 99.8) | 99.3 (98.7, 100.0) | 99.3 (98.9, 100.0) | 99.5 (99.1, 100.0) | 99.6 (99.0, 100.0) | 99.5 (99.2, 100.0) | 99.6 (99.2, 100.0) |
| | | 0.3 | 0.3 | 0.3 | 0.2 | 0.2 | 0.2 | 0.2 |
| Model SE | small | 0.51 (0.39, 0.52) | 0.49 (0.35, 0.49) | 0.48 (0.34, 0.49) | 0.47 (0.32, 0.48) | 0.47 (0.30, 0.47) | 0.46 (0.29, 0.47) | 0.46 (0.28, 0.47) |
| | | 0.6 | 0.7 | 0.7 | 0.7 | 0.7 | 0.7 | 0.7 |
| | medium | 0.45 (0.35, 0.46) | 0.42 (0.31, 0.43) | 0.41 (0.29, 0.42) | 0.40 (0.28, 0.41) | 0.40 (0.26, 0.40) | 0.39 (0.25, 0.40) | 0.39 (0.24, 0.40) |
| | | 0.4 | 0.4 | 0.4 | 0.4 | 0.4 | 0.4 | 0.4 |
| | large | 0.45 (0.37, 0.46) | 0.42 (0.35, 0.43) | 0.42 (0.34, 0.42) | 0.41 (0.33, 0.41) | 0.40 (0.33, 0.41) | 0.40 (0.32, 0.40) | 0.39 (0.32, 0.40) |
| | | 0.4 | 0.4 | 0.4 | 0.4 | 0.4 | 0.4 | 0.4 |
| Relative precision[1] (%) | small | -22.1 (-52.9, -14.8) | -11.4 (-40.6, -8.5) | -9.8 (-28.9, -7.9) | -5.3 (-27.9, -2.0) | -3.2 (-12.6, -1.1) | -1.8 (-7.7, 0.4) | -- |
| | | 2.2 | 2.2 | 2.2 | 2.2 | 2.2 | 2.2 | -- |
| | medium | -26.8 (-51.0, -19.9) | -17.7 (-41.4, -11.4) | -12.9 (-34.3, -7.9) | -9.3 (-18.8, -4.6) | -4.2 (-14.8, -2.5) | -2.3 (-6.0, -1.3) | -- |
| | | 2.2 | 2.2 | 2.2 | 2.2 | 2.2 | 2.2 | -- |
| | large | -24.6 (-29.9, -21.0) | -16.1 (-19.0, -10.5) | -9.7 (-12.0, -5.3) | -6.2 (-8.9, -4.8) | -3.2 (-5.3, -1.7) | -1.6 (-2.9, -0.6) | -- |
| | | 2.2 | 2.2 | 2.2 | 2.2 | 2.2 | 2.2 | -- |
| Mean point estimate | small | -0.05 (-0.07, -0.01) | -0.04 (-0.07, -0.01) | -0.04 (-0.07, -0.02) | -0.04 (-0.08, -0.01) | -0.04 (-0.06, -0.01) | -0.04 (-0.06, -0.01) | -0.04 (-0.06, -0.02) |
| | | -33.2 | -40.3 | -36.7 | -42.0 | -39.1 | -37.6 | -40.1 |
| | medium | -0.73 (-0.76, -0.72) | -0.73 (-0.75, -0.71) | -0.73 (-0.75, -0.71) | -0.72 (-0.75, -0.71) | -0.73 (-0.75, -0.71) | -0.73 (-0.74, -0.71) | -0.72 (-0.74, -0.71) |
| | | -1.9 | -1.8 | -1.8 | -1.8 | -1.7 | -1.7 | -1.7 |
| | large | -1.83 (-1.84, -1.81) | -1.81 (-1.84, -1.80) | -1.81 (-1.83, -1.80) | -1.81 (-1.83, -1.80) | -1.81 (-1.82, -1.79) | -1.81 (-1.82, -1.79) | -1.80 (-1.82, -1.79) |
| | | -0.8 | -0.7 | -0.7 | -0.7 | -0.7 | -0.7 | -0.7 |

1 Negative values represent precision loss. Monte Carlo errors shown are those of variance of $\beta_2$ (source from which Relative precision is calculated).

Table S4: Median values (range) of performance measures for outcome T across all data generating mechanisms, for different sampled comparator (SC) versus full cohort (FC) designs, by effect size. Shaded rows show Monte Carlo standard errors of the median (expressed as %).

| Performance Measure | Effect Size | SC5 | SC10 | SC15 | SC25 | SC50 | SC100 | FC |
|---|---|---|---|---|---|---|---|---|
| Power (%) | small | 10.8 (9.8, 14.7) | 12.8 (11.2, 17.2) | 13.9 (11.0, 17.3) | 14.4 (12.2, 19.3) | 14.8 (12.9, 18.5) | 14.6 (12.3, 19.0) | 15.1 (12.8, 18.9) |
| | | 9.5 | 8.6 | 8.2 | 8.1 | 7.8 | 8.0 | 7.8 |
| | medium | 47.7 (43.4, 79.0) | 54.1 (51.3, 82.3) | 56.8 (52.2, 84.2) | 58.1 (53.1, 85.6) | 59.2 (55.6, 87.0) | 59.9 (56.6, 86.5) | 60.5 (57.2, 86.5) |
| | | 3.3 | 2.9 | 2.8 | 2.7 | 2.6 | 2.6 | 2.6 |
| | large | 84.9 (83.3, 99.7) | 88.6 (87.1, 99.6) | 90.0 (88.3, 99.8) | 91.2 (89.9, 99.8) | 91.8 (90.9, 100.0) | 92.1 (91.1, 100.0) | 92.2 (91.3, 100.0) |
| | | 1.3 | 1.1 | 1.1 | 1.0 | 0.9 | 0.9 | 0.9 |
| Model SE | small | 0.80 (0.51, 0.82) | 0.75 (0.48, 0.77) | 0.73 (0.47, 0.75) | 0.72 (0.47, 0.74) | 0.71 (0.46, 0.73) | 0.71 (0.46, 0.73) | 0.70 (0.45, 0.72) |
| | | 0.8 | 0.9 | 0.9 | 0.9 | 0.9 | 0.9 | 1.0 |
| | medium | 0.64 (0.39, 0.66) | 0.58 (0.35, 0.59) | 0.56 (0.34, 0.57) | 0.54 (0.33, 0.56) | 0.53 (0.32, 0.54) | 0.52 (0.32, 0.54) | 0.52 (0.31, 0.53) |
| | | 0.8 | 0.9 | 0.9 | 1.0 | 1.0 | 1.0 | 1.1 |
| | large | 0.54 (0.33, 0.56) | 0.47 (0.29, 0.48) | 0.44 (0.27, 0.46) | 0.42 (0.26, 0.44) | 0.41 (0.25, 0.42) | 0.40 (0.25, 0.41) | 0.39 (0.24, 0.41) |
| | | 0.7 | 0.7 | 0.7 | 0.7 | 0.8 | 0.8 | 0.8 |
| Relative precision[1] (%) | small | -28.7 (-37.7, -21.9) | -16.9 (-20.9, -12.8) | -10.4 (-14.1, -4.4) | -6.9 (-10.1, -3.4) | -3.3 (-5.7, -1.7) | -1.5 (-3.8, 0.5) | -- |
| | | 2.3 | 2.3 | 2.3 | 2.3 | 2.3 | 2.3 | -- |
| | medium | -35.1 (-39.7, -31.0) | -20.3 (-23.0, -17.8) | -14.0 (-17.4, -11.8) | -8.1 (-10.8, -6.4) | -5.2 (-6.3, -3.9) | -2.1 (-3.3, -1.0) | -- |
| | | 2.2 | 2.2 | 2.3 | 2.2 | 2.3 | 2.2 | -- |
| | large | -45.6 (-51.0, -38.9) | -28.2 (-33.2, -26.7) | -20.7 (-25.2, -17.7) | -14.0 (-16.9, -10.2) | -7.2 (-9.0, -3.8) | -2.9 (-4.9, -1.4) | -- |
| | | 2.2 | 2.2 | 2.2 | 2.2 | 2.2 | 2.2 | -- |
| Mean point estimate | small | 0.42 (0.34, 0.47) | 0.37 (0.30, 0.42) | 0.37 (0.31, 0.41) | 0.36 (0.31, 0.40) | 0.36 (0.29, 0.39) | 0.35 (0.29, 0.39) | 0.35 (0.29, 0.38) |
| | | 5.3 | 5.5 | 5.4 | 5.3 | 5.3 | 5.3 | 5.4 |
| | medium | 1.07 (1.04, 1.08) | 1.04 (1.02, 1.06) | 1.03 (1.00, 1.07) | 1.02 (0.99, 1.07) | 1.02 (0.99, 1.06) | 1.02 (0.99, 1.06) | 1.01 (0.98, 1.06) |
| | | 1.8 | 1.7 | 1.7 | 1.6 | 1.6 | 1.6 | 1.6 |
| | large | 1.61 (1.59, 1.64) | 1.59 (1.55, 1.61) | 1.58 (1.55, 1.61) | 1.58 (1.55, 1.60) | 1.57 (1.55, 1.60) | 1.56 (1.55, 1.60) | 1.56 (1.54, 1.60) |
| | | 1.1 | 1.0 | 0.9 | 0.9 | 0.8 | 0.8 | 0.8 |

1 Negative values represent precision loss. Monte Carlo errors shown are those of variance of $\beta_3$ (source from which Relative precision is calculated).

Table S5: Percentage of models for outcomes L and T where $\beta_2$ and $\beta_3$ could not be estimated, by data model parameters and sampling ratio

| Parameter/sampling ratio | Values | Number of repetitions *N* | Outcome L No estimate obtained for $\beta_2$, *n* (% of *N*) | Outcome T No estimate obtained for $\beta_3$, *n* (% of *N*) |
|---|---|---|---|---|
| Total | | 252 000 | 51 (0.02) | 6093 (2.42) |
| CMB prevalence, $\pi_{CMB}$ | 0.0002 | 63 000 | 15 (0.02) | 2016 (3.20) |
| | 0.001 | 63 000 | 15 (0.02) | 1959 (3.11) |
| | 0.01 | 63 000 | 21 (0.03) | 1495 (2.37) |
| | 0.05 | 63 000 | 0 (0.00) | 623 (0.99) |
| Effect size | Small | 84 000 | 51 (0.06) | 5482 (6.53) |
| | Medium | 84 000 | 0 (0.00) | 585 (0.70) |
| | Large | 84 000 | 0 (0.00) | 26 (0.03) |
| Relative prevalence, $R_{CMB}$ | 5x | 84 000 | 22 (0.03) | 2476 (2.95) |
| | 10x | 84 000 | 7 (0.01) | 2149 (2.56) |
| | 30x | 84 000 | 22 (0.03) | 1468 (1.75) |
| Sampled comparators per case | 5 | 36 000 | 7 (0.02) | 885 (2.46) |
| | 10 | 36 000 | 9 (0.03) | 868 (2.41) |
| | 15 | 36 000 | 7 (0.02) | 868 (2.41) |
| | 25 | 36 000 | 7 (0.02) | 868 (2.41) |
| | 50 | 36 000 | 7 (0.02) | 868 (2.41) |
| | 100 | 36 000 | 7 (0.02) | 868 (2.41) |
| | FC | 36 000 | 7 (0.02) | 868 (2.41) |

FC = full cohort

### Supplementary Figures

Figure S1a: Nested loop plot (power), continuous outcome Y, CHT vs non-CHT, by sampling and DGM

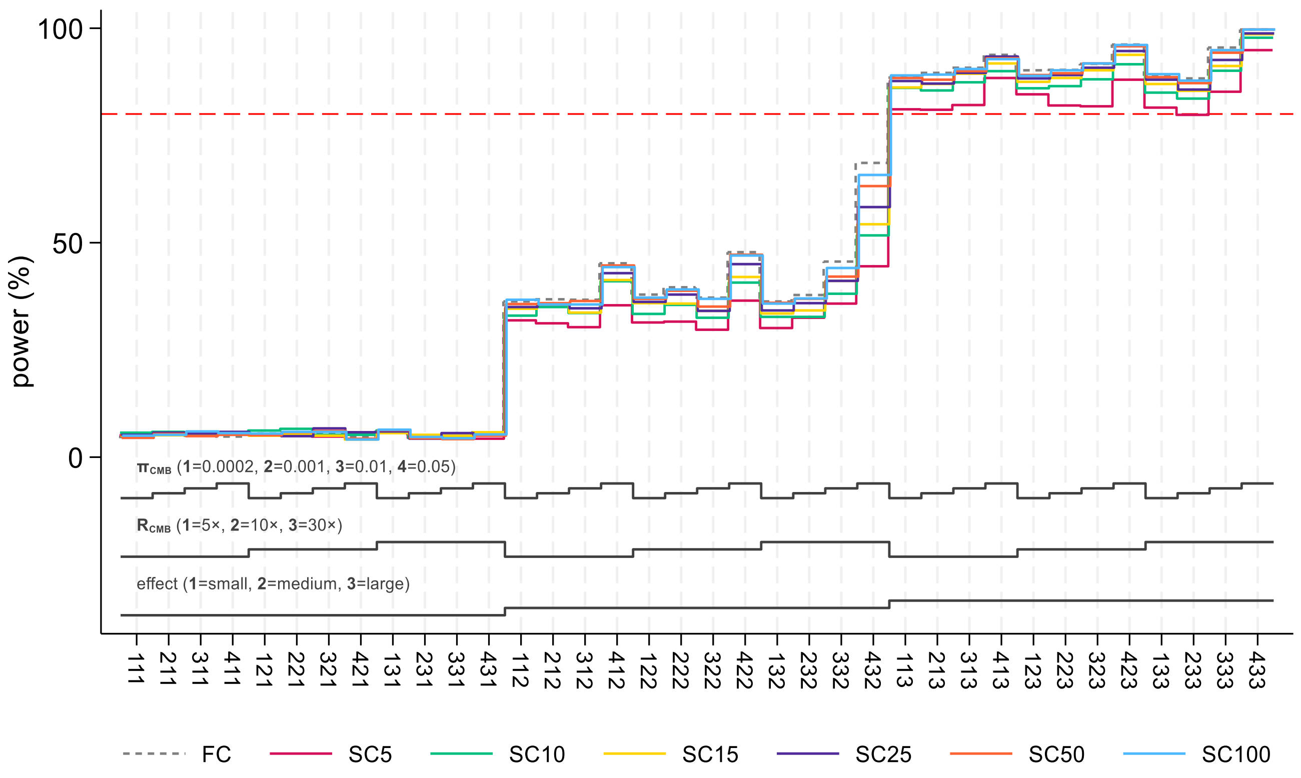


*Red dashed line denotes 80% power*

Figure S1b: Nested loop plot (mean), continuous outcome Y, CHT vs non-CHT, by sampling and DGM

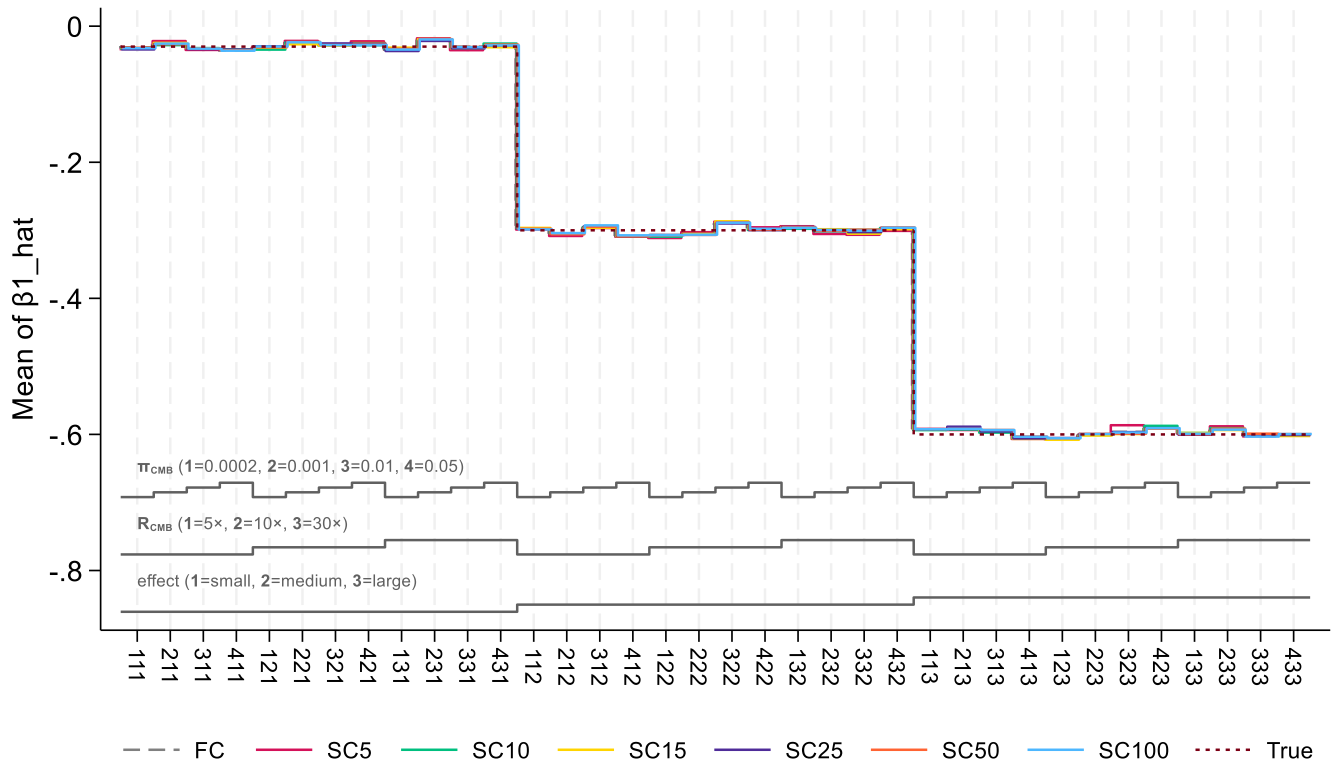

Figure S2a: Nested loop plot (power), binary outcome L, CHT vs non-CHT, by sampling and DGM

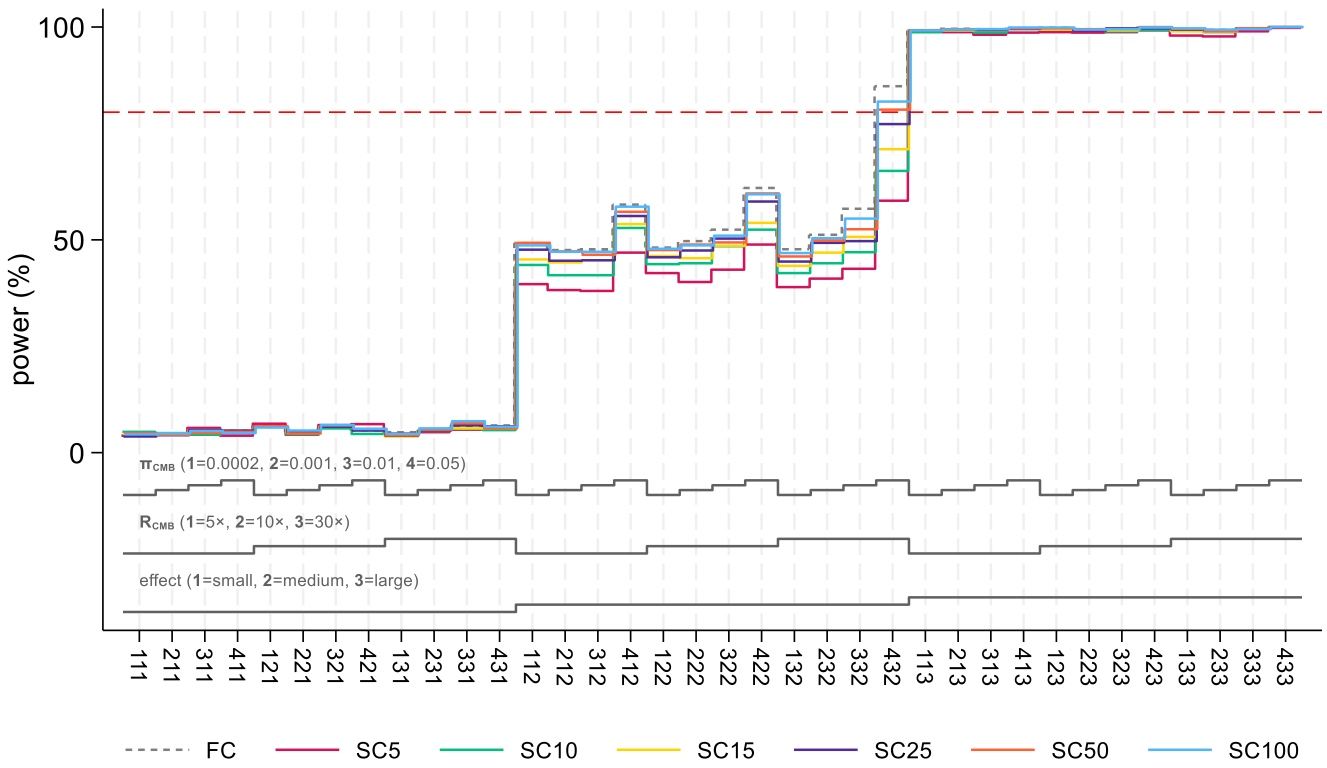


*Red dashed line denotes 80% power*

Figure S2b: Nested loop plot (mean), binary outcome L, CHT vs non-CHT, by sampling and DGM

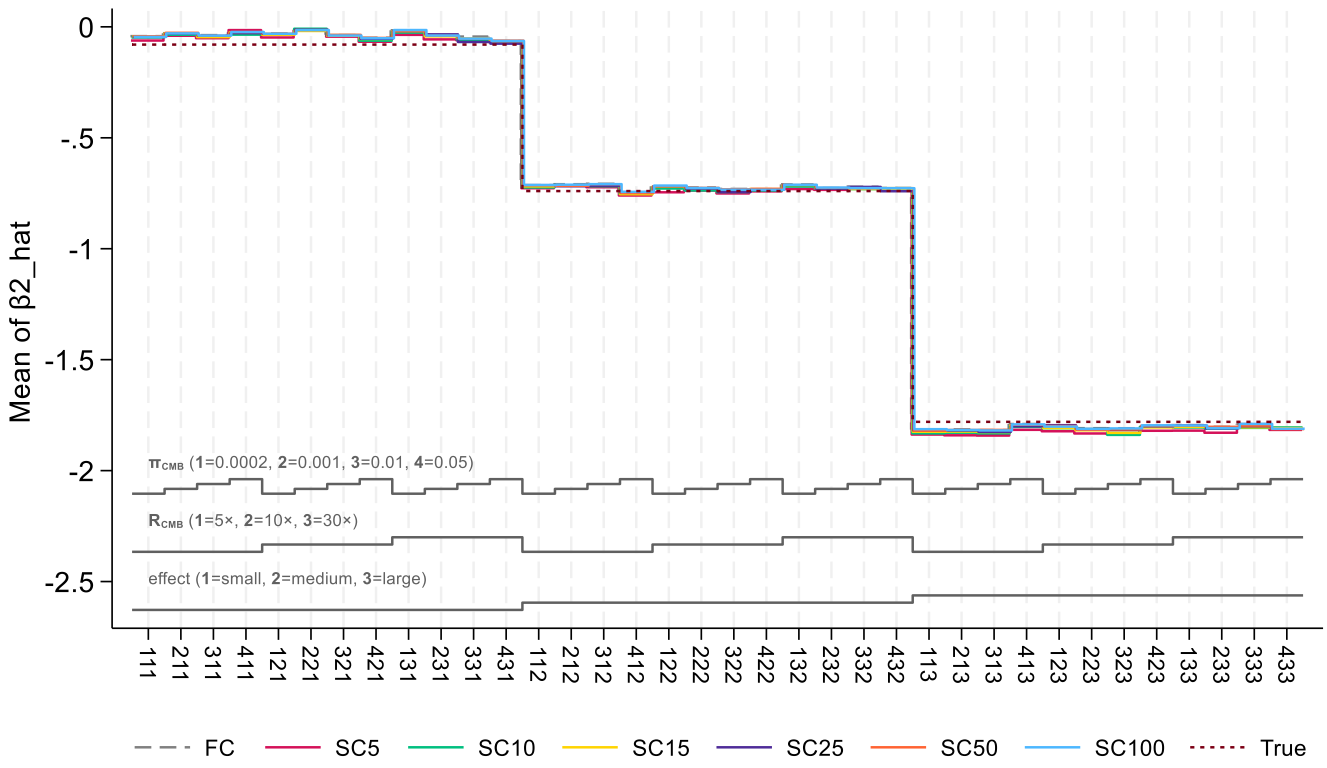

Figure S3a: Nested loop plot (power), survival time T, CHT vs non-CHT, by sampling and DGM

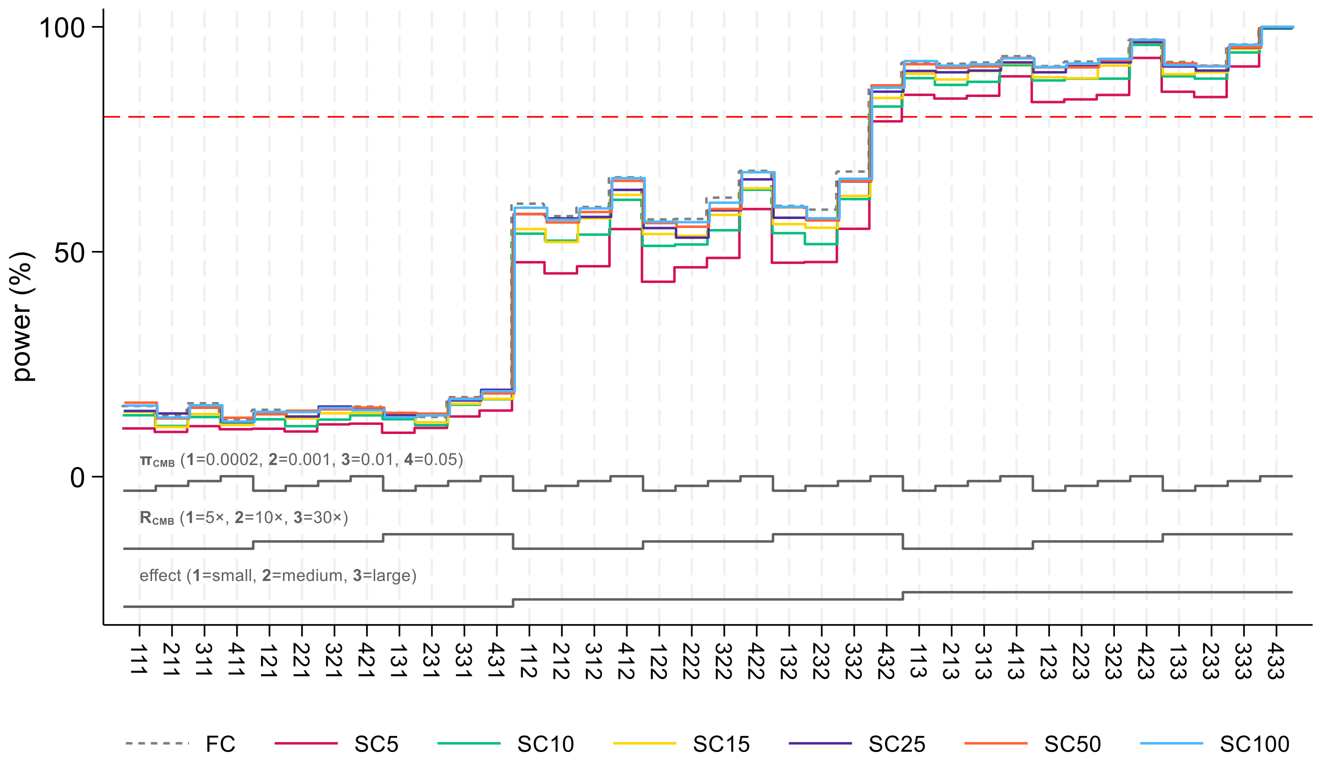


*Red dashed line denotes 80% power*

Figure S3b: Nested loop plot (mean), survival time T, CHT vs non-CHT, by sampling and DGM

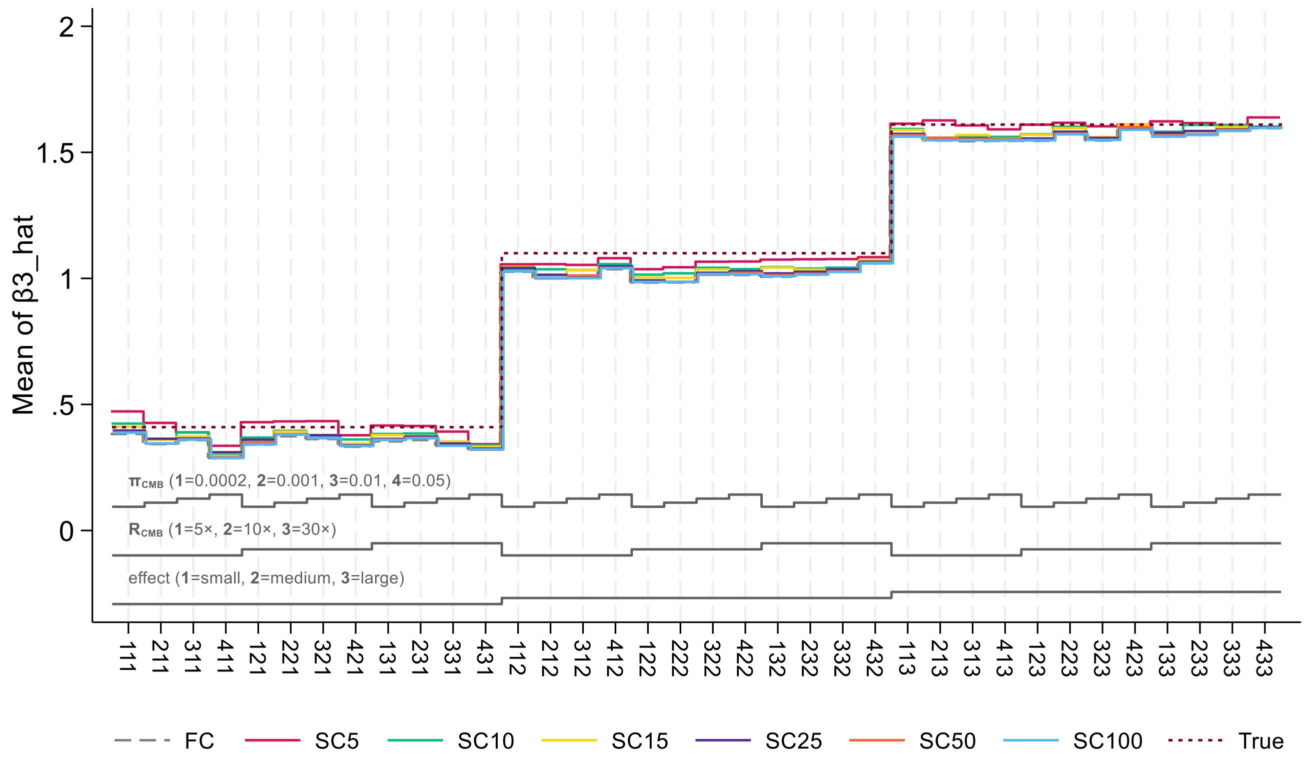
